# Mapping young people’s journeys through mental health services: a prospective longitudinal qualitative study protocol

**DOI:** 10.1101/2022.12.01.22283001

**Authors:** Caitlin Pilbeam, Erin Walsh, Katelyn Barnes, Brett Scholz, Anna Olsen, Louise Stone

## Abstract

Mental ill health is a major health risk for young people. There is unmet need for mental health assessment and treatment across Australia despite significant investment in government-funded plans to cover mental health and youth-oriented services. Understandings of mental health care for young people are impeded by a lack of longitudinal research. Without this research, it is difficult to understand how services do or do not support the recovery of young people over time.

This project will analyse the healthcare journeys of young people aged 16-25 years experiencing their first significant episode of mental ill health, over 12 months in the Australian Capital Territory. The study team will recruit up to 25 diverse young people and their general practitioners (GPs), and conduct four qualitative semi-structured interviews over 12 months with each participant. GP interviews will explore their role in the mental health care and care coordination for the young person. Interviews with young people will explore experiences and perceptions of navigating the health system, and the supports and resources they engaged with during the 12-month period. In between interviews, young people will be asked to keep a record of their mental health care experiences, through their choice of media. Participant-produced materials will also form the basis for interviews, providing stimuli to discuss the lived experience of care.

Through analysing the narratives of both young people and their GPs, the study will establish how young people understand value in mental health care delivery. The study will use longitudinal qualitative mapping of healthcare journeys to identify key barriers and enablers to establishing effective, person-centred health care for young people with mental ill health.

## Introduction

### Young people’s mental health

Australians aged 15-25 experience poor mental health more often and at higher levels than older age groups^1,2^, and 75% of mental disorders that have a life-long impact emerge during this time^3^. Mental health problems cause a high illness burden, particularly for adolescents and young adults^4(pp21-23)^, and elevate risk of self-harm and suicide^5^. Mental ill health can also pose unique challenges for young people, particularly in relation to their employment and education goals and engagement^6,7^.

Some young people are at higher risk for mental ill health and negative outcomes^8^. Risk factors include: low socioeconomic background^9^; history of trauma including sexual, domestic, and family violence^10^; experiences of housing insecurity^9^; unemployment^9^; being female^6(pp4, 10)^; being LGBTQIA+^6(pp5, 10),9,11(p48),12(pp25-33)^; being culturally and linguistically diverse (CALD)^13(pp4-5)^; being Indigenous^6(p5),14(p11),15^; and living in a rural or remote community^16^. Multiple marginalisation can both increase risk of mental health problems, and increase the difficulty of accessing necessary supports.

Between the ages of 15-25 is when many people first develop symptoms of mental ill health, or experience their first diagnosis, and this occurs alongside the life changes and stages of emergent adulthood^17^. For example, individuals may experience shifts in their developmental stage, life conditions, and eligibility for (or access to) particular mental health and wellbeing support services. Care transitions between services or supports occur at different ages depending on the service or health problem^18(pp15, 23)^; segmented and siloed services create significant challenges for navigating the health system^17,19,20^.

### Mental health services for young people

In Australia, young people rely on general practitioners (GPs) more than other mental health services as a source of affordable, accessible, and trusted mental health care, particularly if they experience marginalisation^13(p6)^. Between the ages of 15-19, the proportion of young people who seek support from a GP or health professional increases each year from 44.6% to 60.2%; by age 19, GPs and health professionals are the most preferred source of support for personal issues (preferred by 72.3% of those aged 18-19 years), second only to friends^14(p33)^. Young people also report that GPs provided the most support of any health professional if they needed help coordinating care, and those who received such support reported it as helpful^9^. Still, of those who identified a need for access to mental health care, over a third of 16-24 year olds reported feeling their need for counselling was not fully met^9^.

Despite this apparently central role that GPs play in youth mental health journeys^13(p12),21(p455)^, systemic barriers persist that impede communication between primary care and other services^21(pp659-662)^. These may include bureaucracy, understaffing, limited knowledge and understanding of other services and their offerings, and differing priorities across different organisations^22,23^. In Australia, some services are funded and organised through Federal Government policies and processes, such as General Practice; whereas other services, like hospitals and outpatient units, are funded and managed by State governments. Such differences in funding can further contribute to poor service integration^18(pp21-23)^.

### Mental health care for young people in the Australian Capital Territory

The Australian Capital Territory (ACT) is a small jurisdiction surrounded by another state. Young people living in satellite towns around the ACT may access services both in the ACT and New South Wales (NSW), and via State and Federally funded programs, which presents unique challenges for accessing, co-ordinating and integrating care. The recent Review of Children and Young People in the ACT by the Office of Mental Health and Wellbeing included a project to develop an online portal for young people to assist them in navigating health services^24(p19)^. This “MindMap” tool^25^ offers a “single source of truth” to support young people in self-managing their health service navigation.

In the ACT, young people, carers, and service providers have identified “coping with anxiety and stress” as a critical issue facing young people^24(p9)^. Similarly, just as long waiting times, affordability, and stigma are identified as key obstacles to young people accessing care across Australia^18(p17)^, young people and carers also identify these factors as key obstacles in the ACT^3,24(p12)^.

There has been significant investment in youth-friendly services in the ACT, such as Headspace, Child and Adolescent Mental Health Services (CAMHS), and targeted general practice services (such as The Junction Youth Health Service). Despite this, young people in the ACT continue to struggle to have their mental health needs met^24,26^. Unmet needs are especially concerning for vulnerable young people who experience worse access to care, including Aboriginal and Torres Strait Islander people, people from culturally and linguistically diverse backgrounds, those with disability, and those who are homeless. Availability of services for children and young people with moderate to severe issues is limited^24(p15)^.

There appear to be significant shortcomings in young people’s access to and experience of engaging healthcare services, and there is therefore a need to form a more cohesive picture of the young person’s experience. As yet, we have no research on the experiences of young people with mental ill health as they access and navigate healthcare over time. Such research is critical to better framing mental health care service delivery for young people, and ensuring that bottlenecks and obstructions to access are recognised and addressed.

### Mapping young people’s journeys through mental health care

This study seeks to build an understanding of the healthcare journeys over 12 months of young people aged 16-25 years in the ACT, experiencing their first significant episode of mental ill health. Through analysing the experiences and narratives of both young people and the GPs who work to coordinate their care, the study will establish person-centred value for good mental health care. Through longitudinal qualitative mapping of healthcare journeys, the study will identify key barriers and enablers to establishing effective, person-centred health care for young people with mental ill health. Overall, this research will identify and describe the needs of young people accessing mental health services, and how the ACT healthcare system can best provide equitable, timely, and high-value care.

This project will generate:

1. A summary set of key barriers and enablers to effective service provision at different points in the young person’s journey through the mental health system, and potential routes to overcome or circumvent these barriers for healthcare providers.
2. A set of recommendations for key determinants for assertive outreach for young people based on their own experiences; and in-reach models that enable better collaboration between GPs and specialist mental health services.
3. An articulation of patient-determined values for mental health care for young people, and assessment of how and when this was met in the participant’s journey.
4. A tool for young people using graphic feedback methods: a multi-page story describing strategies and approaches used by young people navigating the health system, and the resources they draw upon in self-management and support through that time.

## Methods & Analysis

### Aims & Research Questions

Using in-depth, longitudinal, narrative and creative approaches, the study aims to explore diverse patient and clinician experiences, amplify the voices of young people facing mental ill health to support greater advocacy for their needs, and to identify potential areas of improvement for health services in the ACT. Findings will be used to inform health service and system planning to support appropriate, affordable and streamlined access to mental health services for young people.

This study is being conducted by an interdisciplinary team whose complementary experience and expertise spans qualitative research, education, health services design, clinical health service provision, health policy, and science communication. The team includes a GP, child psychiatrist, dietitian, medical anthropologist, critical health psychologist, youth worker, social worker, and a graphic artist. Together, the qualitative and creative methods and approaches we will employ throughout the study will provide a holistic, grounded, real-time understanding of young people’s mental health care journeys.

The study will explore the following research questions:

1. How do young people experiencing their first episode of significant mental ill-health navigate the journey through mental health care?
2. What are the human resources for support that young people draw on in that time?
3. What are key barriers and enablers to accessing effective care for young persons?
4. What are the experiences of GPs in supporting the navigation of young persons through health care to receive effective mental health care?
5. What are the key values of good health care for persons experiencing their first bout of mental ill health?

### Study Design

We will undertake a prospective longitudinal qualitative study over 12 months of young people with symptoms of mental ill health, and their GPs. We will map each participant’s patient journey from their own perspective, and the perspective of their GPs to form an understanding of subjective participant experience in relation to health service engagement and objective support offered. Prospective longitudinal design is the gold standard for patient journey mapping, as it limits recall bias, and allows for an in-depth and evolving understanding of patient experience ^27^. Prospective longitudinal qualitative designs have previously been successful in exploring young people’s experiences of seeking and receiving health care ^13^.

A flowchart showing the study design is given in **Figure 1**.

**Figure 1:**
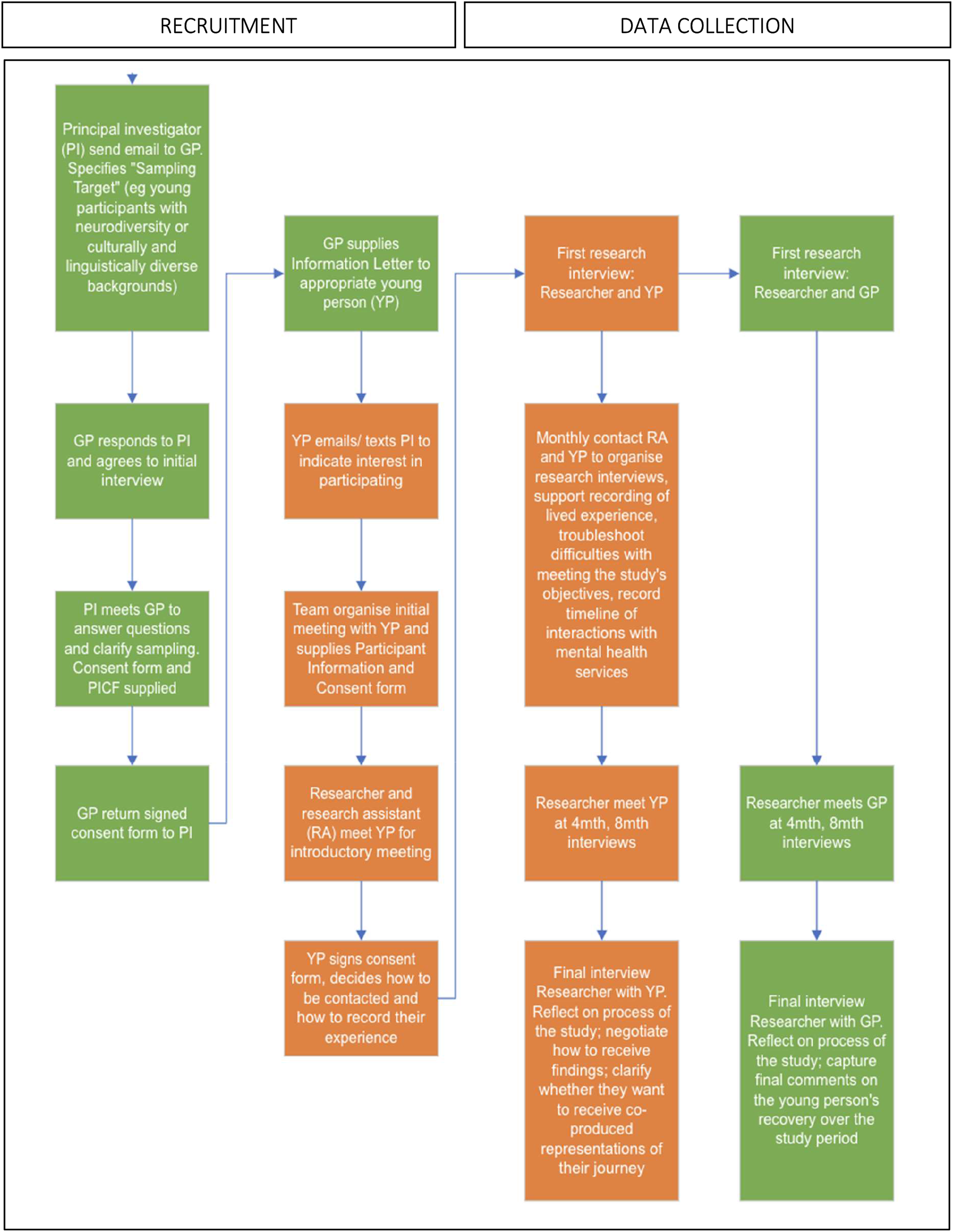
**Study design flowchart** showing research activities with GP participants in green and journey participants in orange.

### Research steering groups

A Steering Committee and a Young Person’s Research Panel will inform the design and conduct of this study. The Steering Committee and Research Panel will meet separately four to five times over the study duration. The purpose of these groups is to collaboratively refine recruitment and data collection methods, inform the interpretation of data, and develop a dissemination strategy for key findings to policy, healthcare practitioners, and young people.

The Steering Committee will provide critical insights into the clinical, health service design, and policy environment in which the study occurs. The Young Persons’ Research Panel will ensure that the study recruits and supports appropriate young people in the study; that the research methods are appropriate to the cohort; and that the findings are interpreted and translated in a way that meets the needs of young people in the ACT.

#### Steering Committee

A group of up to 15 experts in youth mental health and representatives from a range of relevant local, national, and government organisations. Experts were identified by the research team through stakeholder analysis and includes representatives with expertise in clinical, policy, local and national mental health. Members who are not supported by their organisations will be paid for their time, travel, and opportunity cost of participation.

#### Young Person’s Research Panel

A group of up to 12 young people aged 16-25 years, who may have had mental ill health themselves or been affected by peers’ mental ill health. Members will be recruited through schools, universities, GP surgeries, and community organisations, and paid for their time, travel, and opportunity cost of participation.

### Participants

#### Journey participants

Young people aged 16-25 years with high-prevalence mental health conditions (e.g. depression, severe anxiety, eating disorders), who have accessed or attempted to access mental health services (e.g. public or private psychology or counselling services, public or private psychiatry) in the three months prior to first contact, currently living in the ACT or surrounding NSW area. Participants will be excluded if they have acute psychosis at intake interview, participation is deemed to be unsafe, or they cannot provide consent.

GP practices will be purposively sampled to represent diverse populations, and individual GPs will be asked to invite young people who fit the study criteria to consider participating in the research. We aim to recruit between 16 and 25 young people. While we do not seek to be representative, we wish to identify a diversity of experiences from which to learn. Given the longitudinal nature of the study, this number is also what is practicable for the researchers whilst maintaining rigour and participant engagement.

#### GP Participants

Any GP that journey participants visit with regards to mental health services and support, in the ACT or surrounding NSW area.

### Recruitment and Consent

GPs will be informed of the upcoming study through local practice-based research and interest groups, and General Practice newsletters. GPs will be approached individually according to a theoretical sampling model ^28^ to achieve maximum diversity. It will be made clear that GPs will be remunerated for their time, based on Medicare rebate rates.

Journey participants will be recruited from general practices using a diversity sampling frame to capture breadth of backgrounds. GPs who have expressed interest in the study will identify eligible young people from their current patient list, and mail these potential journey participants study invitation letters. GPs are well positioned to assess capacity of the young person to understand and consent to research, and capacity to participate in an interview. The young person can then contact the research team by text or email if they wish to participate.

Providing young people with information about the study is at the GP’s discretion. The research team will not be informed of young people who been invited to participate. It will be made clear to all invited young people that not participating in this study will not influence their current treatment or relationship with their treating health professional.

When a potential journey participant contacts the research team, a research team member will explain the research, discuss study involvement, and provide any further information. It will be made clear that they will be remunerated for their time $100 at the end of each interview (and transport costs remunerated). This aligns with similar remuneration for Jury Duty, also a civic service, and recognises the preparatory work participants undertake prior to interviews. The researcher will ask participants if they wish for a parent or carer to be present to explain, interpret, or assist with an interview (in line with the NHMRC (2018) National Statement on the Ethical Conduct for Human Research (NSECHR) standard 4.3.2)^29^. If the participant consents to parent or carer presence, then verbal consent of the parent or carer will be sought at the interview.

Parental consent will not be sought for journey participants taking part in the study. A minor may consent to treatment without prior consent of the parent, if they are deemed by their clinician to be Gillick competent^30^. In these circumstances, parents may not be aware of their child’s mental health or their treatment. If a young person is able to understand and consent to treatment for mental health, their experiences of accessing and navigating mental health services are valid and worthy of being explored despite parent knowledge or awareness of said mental health treatment (in line with section 4.5.3 of the NHMRC (2018) NSECHR)^29^. With the participant’s permission, parents or carers are able to contact the research team to ask questions about the study methodology, rationale, and purpose. Parents or carers not present at an interview will not receive information about individual participant responses to interview questions.

### Data Collection

#### From GPs

For each journey participant, the treating GP will be asked to participate in a set of four interviews over 12 months. These interviews will explore the journey of the young person through the mental health system. This will provide context regarding the health system and navigation that may not be immediately apparent to the young person involved (e.g. successful and unsuccessful referral pathways). Interviews will include discussion of choice of referral, and will explore why GPs chose, or did not choose, certain referral pathways.

#### From journey participants

Young people participating will be followed up over a period of 12 months. We will conduct four semi-structured qualitative interviews with each participant; either every four months, or as identified by the participant. Research team members will be trained to conduct interviews and keep fieldnotes of all participant interactions^31,32^. Interview one will explore how the participant initiated help seeking (i.e. becoming concerned about mental health; initial information-gathering; requests for support; actions taken by care-providers). Interviews two, three, and four will explore the participants’ experiences of navigating the health system, and what makes this harder or easier for them. Interview four will also engage with reflection on experiences over the last 12 months. Participants will have the opportunity to review their transcripts and add to their reflections for four weeks after each interview.

In between interviews, each journey participant will be asked to keep a record of their experiences seeking and accessing mental health care, in the way most suited to them. These participant-produced materials will serve as a memory-aid and, in concert with the semi-structured interview guide, will provide stimuli to discuss the lived experience of care in alignment with the issues most salient to the young person. To reduce attrition and support ongoing record-keeping, the research assistant will follow-up remotely with each participant every month. Participants will be given the choice of keeping these records via arts-based methods.

#### Participant-generated reflective data

Arts-based methods are theoretically grounded in ethnography, anthropology, sociology, psychology, and arts-based research^33–36^. Involving a wide array of approaches, arts-based methods can be broadly described as forms of data collection where participants may curate, create, or respond to imagery or other forms of creative expression (e.g. select from childhood photos; take new photographs articulating their lived experiences; respond to photographs provided by the researcher).

These methods include a wide range of mediums, from dance to sculpture, though the most common are photography and drawing (as used in autophotography, photovoice, visual diaries, and draw-and-write). This suite of arts-based methods reduces reliance on spoken data and participants’ ability to articulate lived experience^37^; is appealing and interesting to participants; and centres and empowers participant voices by providing them greater agency in how they engage with the researcher^33,38^. At each interview, participants will have the opportunity to choose how their personally-generated materials are used by researchers.

### Data Analysis

Given the blend of qualitative methods, including interviews and arts-based methods, data will be a combination of: (1) interview transcripts; (2) fieldnotes; and (3) participant-generated materials. Utilising these different sources and forms of data uniquely offers a more holistic picture of lived experiences and journeys over time than one data source would alone. In order to maintain the nuance of these data, we will adopt an iterative approach throughout analysis that allows us to bring these different data together, informed by narrative^39,40^ and thematic analysis^41^.

We will utilise the principles of the pen portrait method^42^ to create ‘case summaries’ of each participant’s longitudinal narrative, drawing on the different data sources to populate them, and GP’s accounts and chronology to further contextualise. This will involve identifying elements of narrative (e.g. event-challenge-overcome), story-telling devices used to describe the journey, and strategies used to address challenges. Thematic analysis of interview transcripts in tandem with this process will further develop the summaries. We will then compare and contrast these summaries, ‘zooming in’ and ‘zooming out’^43^ to explore individual differences and commonalities to experiences, events, structure, etc.

Interview transcripts will be analysed using the six phases of thematic analysis: (i) familiarising with the data; (ii) generating initial codes; (iii) searching for themes; (iv) reviewing themes; (v) defining and naming themes; and (vi) producing the report. The research team will use a systematic and iterative approach to understanding the meaning of phenomena within the specific context of the participant’s journey. Thematic analysis will also be used to identify general themes relating to young person and GP experiences navigating mental health services.

### Ethical Considerations

The study was approved by ACT Health Human Research Ethics Committee on 24 February 2022 (2022/ETH00014). The research team acknowledges the complex ethical challenges in targeting young people experiencing mental ill health for research, due to their vulnerability in terms of age and health. In line with section 4.2 and 4.5 of the NSECHR, to minimise the risk of coercion, discomfort, and distress the research team will:

- Have a GP external to the research team, with knowledge of the patient, judge the competence of the patient for study participation. Any GP that takes part in recruitment and interviews will be provided with information on judging competence to take part in an interview.
- Have researchers with experience in adolescent mental health and qualitative methods explain the research and conduct interviews with young people and/or carers. These researchers will be able to detect distress, determine competence, and will respond accordingly via ending the interview, advising on support services, and communicating back to the nominated treating GP.
- Use information statements written in plain language, at a year 7 reading level or below.
- Involve patient parents or carers, when appropriate and with patient consent, to help interpret and explain information.
- Explain that interviews can be paused, terminated early, and/or restarted another time if requested.
- Explain that participants are free to withdraw consent at any time with no impact on their care or their relationship with any stakeholders involved.
- Explain that information provided will be de-identified, and that only the interviewer will be able to link responses to a participant. The only purpose of the interviewer being able to link an individual to their responses is to remove responses if consent is withdrawn.
- Interviewers will undertake purpose-developed training to conduct interviews with high methodological rigor while also maintaining participant health and wellbeing.
- Interviewers will debrief with senior members of the research team, maintaining confidentiality, as part of maintaining rigour and the safety of all those involved.

## Discussion

Research into youth mental health is crucial for identifying gaps in service provision, challenges to access and use, and unmet needs. Cross-sectional research captures usage data, but cannot capture the decision-making processes; in particular, decisions to avoid or disengage from services. Our study addresses these aspects by mapping mental health care journeys as they unfold over a year, from the perspective of both the young person seeking support and the GP coordinating their care. We fully anticipate that these ‘journeys’ for individual participants may be multiple and may not conform to a linear narrative. Our innovative use of a combined qualitative longitudinal approach, involving repeat interviews complemented by fieldnotes and creative arts-based methods, allows us to collect rich and detailed data that speaks to different facets of participants’ experiences and shifts in experience over time.

Alongside interviews, fieldnotes are a medium through which researchers can immerse themselves in research contexts and interactions with participants, attending to that which may not be explicit in written or spoken form, and thus supports interpretation of findings^31,32^. Further, nonverbal approaches allow for non-linearity in meaning construction, making creative arts-based methods useful for the articulation of painful or traumatic experiences, which are frequently not perceived sequentially^37^. They are similarly helpful for those who struggle with verbal expression^34,35,37^.

Over-researched groups, or people who need to repeatedly share their internal state (as occurs in accessing mental health services), could supply data that is diluted by the development of a narrative refined by repeated verbal or written articulation of their experience. Arts-based methods can “make the familiar strange” by shifting discourse from the verbal to the visual and creative, which in turn can reveal fresh insights^44^. Additionally, once research is complete, the materials (e.g. images) created during the research process can serve as an accessible way to convey results^35,38^.

### Amendments: Steering Group and Young Person’s Research Panel

A key component of developing a research approach and questions that are relevant, appropriate, and valuable to the stakeholders of this research area has been the incorporation of the Steering Group and Young Person’s Research Panel^45^. We have so far collaboratively refined our research protocol with both these groups, which have each met twice to date. These meetings involved and will continue to utilise an iterative process of discussing the research design, gathering suggestions, and feeding back any changes made. This process ensures that participants in each group understand how they have been heard and heeded.

We are engaging the diverse expertise of these groups – in the lived experience of accessing and provisioning youth mental health services – particularly to enhance recruitment, retention, the relevance of methodology and interview questions to young people and GPs, further inform interpretation of data, and provide input to enhance dissemination of findings^45–47^. In the event further potential amendments and incorporating suggestions, in future, we will continue the same process in consultation with these two groups of key stakeholders at the scheduled meetings throughout the study.

### Dissemination of Findings

The research team has identified multiple avenues and formats for disseminating research findings to diverse audiences, including academic, policy, and public outputs. The authors will take precautions to ensure no individual is identifiable in reports or publications. We anticipate that the findings from this study may be used to inform mental health services and policy in the ACT, and to inform future research projects centred on mental health services for young people in the ACT and comparable settings. As above, findings dissemination will be further shaped by consultation with the Steering Group and Young Person’s Research Panel.

Outputs will include:

- Graphic materials, led by the member of a research team who is also a graphic artist, and supported by the young persons’ research group, communicating study findings to a general audience.
- Scholarly journal articles and presentations shared at relevant health and policy conferences.
- Full text scholarly manuscripts produced from this study will be shared with participants upon request.
- De-identified aggregate findings to be reported to the funder and local mental health policy and interest groups and organisations.
- A plain language summary of the de-identified aggregate results to be shared with all participants.
- Co-constructed ‘maps’ for journey participants to take away with them and use as they wish.

### Challenges and Limitations

We recognise that the study presented has multiple challenges and limitations that need to be addressed across the research process, to mitigate potential biases and ensure appropriate interpretation of findings. The study:

- Relies on GP engagement for sampling, meaning that the research team loses some control over targeting sampling.
- Has a small sample size which, combined with the mental ill health burden of participants, may contribute to high risk of dropout. While dropout is a datapoint in itself, it may mean less data can be collected overall.
- Multiple methods yield rich and strong insight, yet may be more difficult to synthesise concisely and coherently. While rich data is a strength as this reflects the nature of experience, it can hinder research translation if simplicity and precision is preferred.
- Generalisability is limited to mild-to-moderate mental ill health as participants need to be well enough to participate over 12 months. The timeframe also precludes engagement with young people who are itinerant.
- Whilst the research team will seek to minimise the study’s impact on participants, we are aware that it may be impossible to fully disentangle the therapeutic impact of self-monitoring and reflection that participants undertake during data collection.
- Similarly, GP’s decision-making may be impacted by knowing that these decisions will be analysed by the research team and in a different way than usual.

## Supporting information

Supporting Information: Interview Schedules

## Data Availability

Data not available due to ethical restrictions.

## Author Contributions

***Caitlin Pilbeam*** – Conceptualisation; methodology; project administration; investigation; writing

– original draft preparation; writing – review & editing

***Erin Walsh*** – Conceptualisation; methodology; project administration; investigation; writing – original draft preparation; writing – review & editing

***Katelyn Barnes*** – Conceptualisation; funding acquisition; methodology; investigation; writing – review & editing

***Brett Scholz –*** Conceptualisation; funding acquisition; methodology; writing – review & editing

***Anna Olsen*** – Conceptualisation; funding acquisition; methodology; supervision; writing – review & editing

***Louise Stone*** – Conceptualisation; funding acquisition; methodology; project administration; investigation; supervision; writing – review & editing

## Acknowledgements

We recognise and value the contribution of the Steering Group and Young Persons’ Research Panel in guiding the research team.

