## Supporting Information: Interview Schedules for "Mapping young people’s journeys through mental health services: a prospective longitudinal qualitative study protocol"

INTERVIEW SCHEDULE FOR GENERAL PRACTITIONERS

<u>Interview #1</u>

1. **What are your concerns about this young person?**
2. **Who are you planning to involve in their care and why?**
3. **Are there any barriers to treatment that you see? How do they affect what you do and where you direct your referrals?**
4. **What follow-up are you planning to offer and why?**
5. **Can you tell me about your experience of caring for young people in the ACT? What do you see as the enablers and barriers to care? What is your personal experience of caring for young people like?**

<u>Interviews #2 and #3</u>

1. **Has your understanding of this young person's needs changed over time?**
2. **What do you think has been their experience of help seeking? How have you assisted them in finding the help they need?**
3. **Do they have unmet needs that you would like to address? What are the barriers to getting those needs met?**

   *Probes:*

   How do you know about those needs?
4. **Are there areas where your goals and their goals don't align? How do you manage this?**
5. **What has been your experience of caring for this young person over the last four months?**

<u>Interview #4</u>

1. **How would you describe this young person's journey around the mental health care system?**
2. **What have been some of the benefits and some of the challenges?**
3. **Have there been needs that have not been met?**
4. **What has been your role in their care in the last 12 months?**
5. **Have there been issues in coordinating care for this young person?**
6. **What services would have smoothed the recovery journey for this young person?**

INTERVIEW SCHEDULE FOR JOURNEY PARTICIPANTS

Interview #1

1. **Before we start, could you tell me a bit about yourself?**
   *Probes:*
   What do you spend your time doing?
   What is important to you in your life?
   What was it about this research project that interested you?

2. **You're part of this project because you have some experiences accessing health services for your mental health. Can you tell me a bit about how that journey started?**
   *Probes:*
   How did you first realise you might need / wanted some support?
   Did something happen? Did someone say something to you?
   Did you book any appointments? Did anyone help you do that?

   a. **Who did you first talk to about your mental health?**
      *Probes:*
      Or did someone talk to you?
      What was that conversation like for you?
      What did you do next?

   b. **When you were first finding out about your mental health, did you access any online resources?**
      *Probes:*
      What did you find helpful? Or not helpful?
      What was it like using those resources (e.g. website, app)?

3. **How did your GP become involved in your mental health?** (Reminder that this is confidential and won't be shared with your GP.)
   *Probes*:
   How did you first start talking to them about mental health? Who raised it?
   Did anyone help you with that? Who? How did they help you? When was this?
   Did you choose that GP specifically? How did you choose them / feel about them?
   How did you feel talking about it with them?
   Are they still involved in your mental health care now? How is that going?

4. **Has your GP made any referrals to other health services for you?**
   *Probes:*
   Do you know where the GP has referred you? Did you know about this service before?
   Have you been? How did that go?
   How do you feel about the referral(s)?
   Did you ask for a referral? To somewhere specific? How did you choose?
   Is there any kind of care you specifically want? How do you know about it?
   What do you think the next steps are going to be for your mental health care?

5. **Aside from your GP, have you been to see any other healthcare professionals about your mental health?**
   *Probes:*
   How did that go?
   Did anyone help you with that? How did you find / hear about them?

How did you feel?
How did the healthcare professional respond?

6. **Last time we met, we talked about how you'd record your experiences and share them with us. What did you decide to do?**
   *Probes:*
   How has that been going over the last few weeks?
   Have you got what you need to do that?
   What sorts of things have you created?

   a. **Would you like to go through some of the things you've created? You can talk me through them.**

7. **This research is about how young people navigate the mental health care system. Looking back on your mental health care journey so far, has anything made it easier for you to get mental health care?**
   *Probes:*
   Has anything made it harder?
   Has anything surprised you about the process so far?
   Have you had support from people around you?

   a. **Are you using any other services to support your mental health?**
      *Probes:*
      Online or face-to-face?
      People, programmes, **apps**, places that you go that help you? (e.g. Vinnies, Headspace)

8. **Before we finish, is there anything else you'd like to say or add that we didn't get a chance to talk about yet?**

   a. **Are there any other questions that you think we should be asking people in this study?**

<u>Interviews #2 and #3</u>

1. **How is recording your experiences going?**
   *Probes:*
   Have you got what you need to do that?
   What sorts of things have you created?

   a. **Would you like to go through some of the things you've created?**

2. **Last time we talked about your first GP appointment where you discussed your mental health. Have you had any GP appointments in the last few months?**
   *Probes:*
   How did that go? How did you feel?
   Who made the appointment?
   Did anyone come with you?

3. **Have you had any mental health appointments with <u>other</u> professionals?**
   *Probes:*

What kinds of health professionals (e.g. counsellor, psychologist, psychiatrist)?
How many times have you met with them?
How did you come to see them? How did you know about them?
Who made the appointment? Did anyone come with you?
How has that been going?

a. **Were there any appointments that you didn't go to?**
*Probes:*
What made you decide not to go?

b. **Have you been <u>back</u> to see [insert healthcare professional]?**
*Probes:*
How did that go? How did you feel?
When was that? How many times did you see them?
How did the healthcare professional respond?

c. **Have you been to any new services, or accessed any online resources, even if you didn't speak to a healthcare professional about your mental health?**
*Probes:*
How did that go? How did you feel? Did you find it helpful?
What made you decide to access those resources?
How did you hear about them?
Was anyone with you?
When did you go? How many times? Do you think you'll go back?

4. **Do you have any appointments coming up in the next few months?**
*Probes:*
Who with? The same or different healthcare professionals?
How do you know about them? Who made the appointment

5. **Looking back on your mental health care journey so far, what things do you think have made it easier for you to get mental health care?**
*Probes:*
What has made it harder?
Has anything surprised you about the process so far?
6. **Before we finish, is there anything else you'd like to say or add that we didn't get a chance to talk about yet?**

a. **Are there any other questions that you think we should be asking?**

<u>Interview #4</u>

1. **Thinking back over the last 12 months of your mental health journey, how did you feel at the beginning?**

a. **How do you feel now?**

2. **Who have been the most important people along the way?**
*Probes:*
Who in your life has supported you to get (the right) care for your mental health?
How have they supported you? When?

3. **Thinking about the mental healthcare appointments you had, what would you advise other young people to do in similar situations?**
   *Probes:*
   What made things easier for you to get the care you needed?
   What made things harder?
   What surprised you along your journey?

4. **How has accessing mental health care impacted your life more broadly?**
   *Probes:*
   What has helped you manage your mental health?
   What has not helped?
   What were the indirect benefits of accessing mental health care? (E.g. social life)
   What were the indirect challenges? (E.g. on finances)

5. **Thinking about the different mental health appointments you have had, what were some of the best things that happened?**

   a. **What were some of the less great things that happened?**
      *Probes:*
      What could have been done differently to improve your experience?
      What message would you give any of the people or services you engaged with?
      What do you want them to know about what was / wasn't helpful?
